# Calibrating an Epidemic Compartment Model to Seroprevalence Survey Data

**DOI:** 10.1101/2020.05.27.20110478

**Authors:** Michael Halem

**Affiliations:** BeCare Link LLC; Rocket Power LLC

**Keywords:** Seroprevalence, model, calibration, SIR, compartment, epidemiology

## Abstract

To date, the Covid-19 epidemic has produced tremendous cost and harm. However, to date, many epidemic models are not calibrated to seroprevalence survey(s). This paper calibrates a relatively simple, SIR plus confirmed cases (“SIRX”) model against seroprevalence survey data released by the State of New York. The intention of this paper is to demonstrate a potentially new technique of calibration for epidemic models used by scientists, public health officials and governments. The technique can then be incorporated in other more complex models. Open source code is included to assist model developers.

## Introduction

The intention of this paper is to provide a calibration technique, applied to a relatively simple SIR plus cases model (“SIRX”) using seroprevalence data. This paper is an attempt to communicate to epidemiologists and other modelers within a timeframe that is useful to managing the current epidemic. Numerous simplifications are made to concentrate this communication. Hence additional detail and accuracy are explicitly beyond the scope of this paper.

All code and data are available at [1]. This calibration technique was independently developed by the author. An extensive search for other similar techniques which may have existed in the literature was beyond the time scope of this paper. Such techniques will be credited in subsequent revisions of this paper as they come to the attention of the author.

### Generalizing an Antibody Test

As will be explained in detail below, in this paper the New York State “Wadsworth” Antibody Seroprevalence Survey will be used to calibrate a SIRX model.

In the attached appendix, a simplified antibody test is used to demonstrate via linear algebraic manipulations how both sensitivity and specificity would interact with different (perfectly) known seroprevalence time series, assuming that the test is sampled at a fixed time relative to the time series independent variable t. Antibody test sensitivity and specificity curves were rounded from the Wadsworth test in the Appendix so that the arithmetic can be checked by inspection. In the Appendix, and summarized in Figures 1, 2, and 3 below, three different assumed time series are put forward as examples: Figure 1: a cumulative infection growth rate doubling every 3 days (newly infected doubling every 3.64 days), representing an epidemic’s typical exponential growth before the introduction of social distancing measures; Figure 2: a flat (constant) infection rate per day; and Figure 3: an exponential decrease in the number of new infections at the same 3.64 halving rate as Figure 1, created by “playing backwards time” from the example Figure 1.

**Figure 1.**
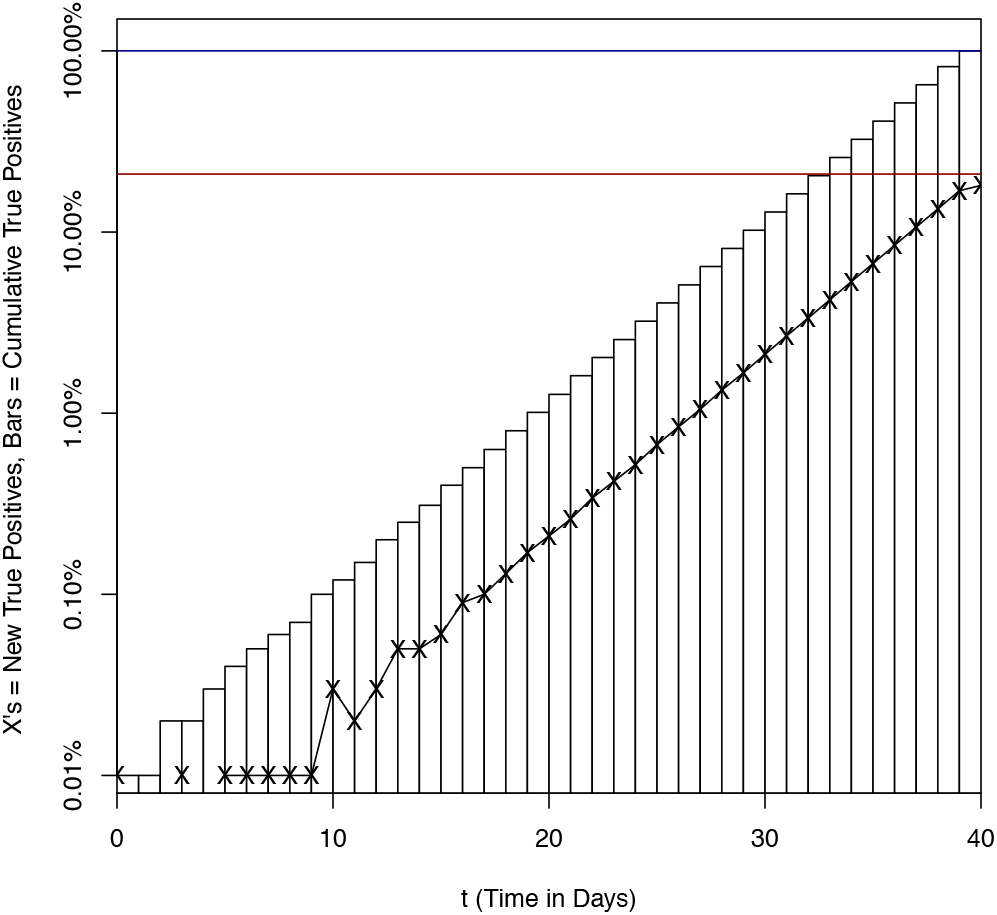
Exponential Increase of New Infected Time Series

**Figure 2.**
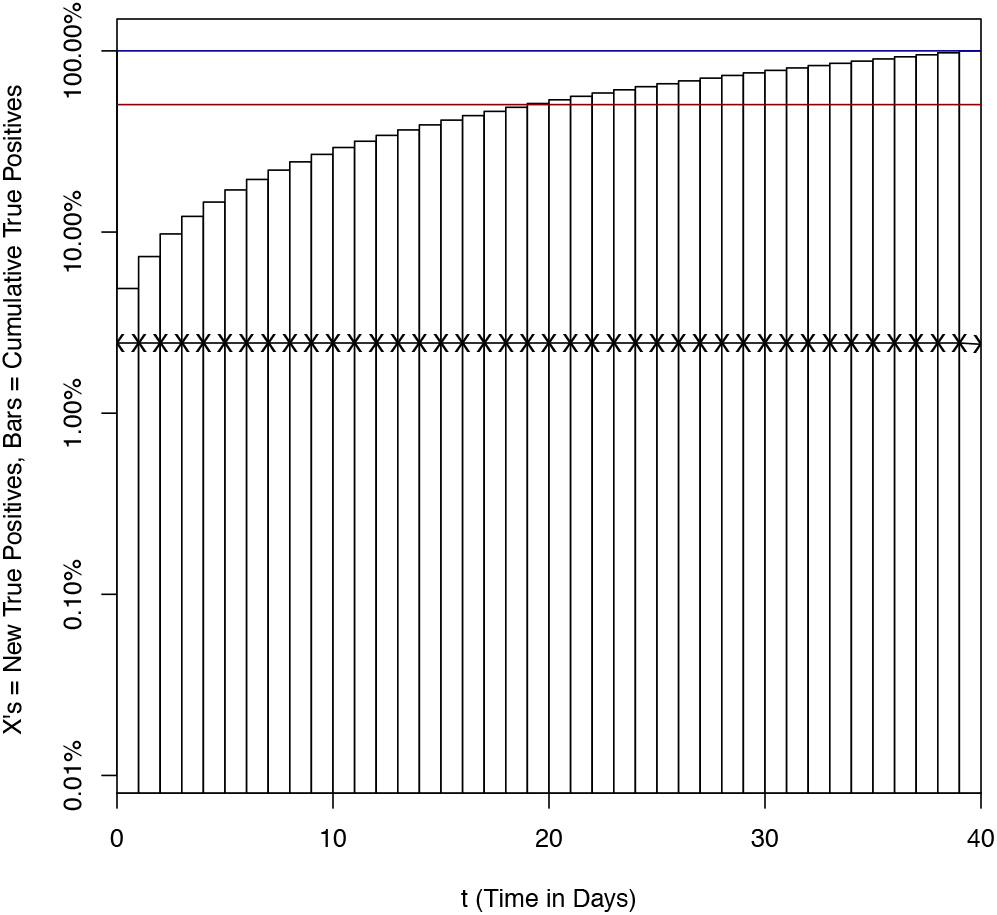
Flat New Infected Time Series

**Figure 3.**
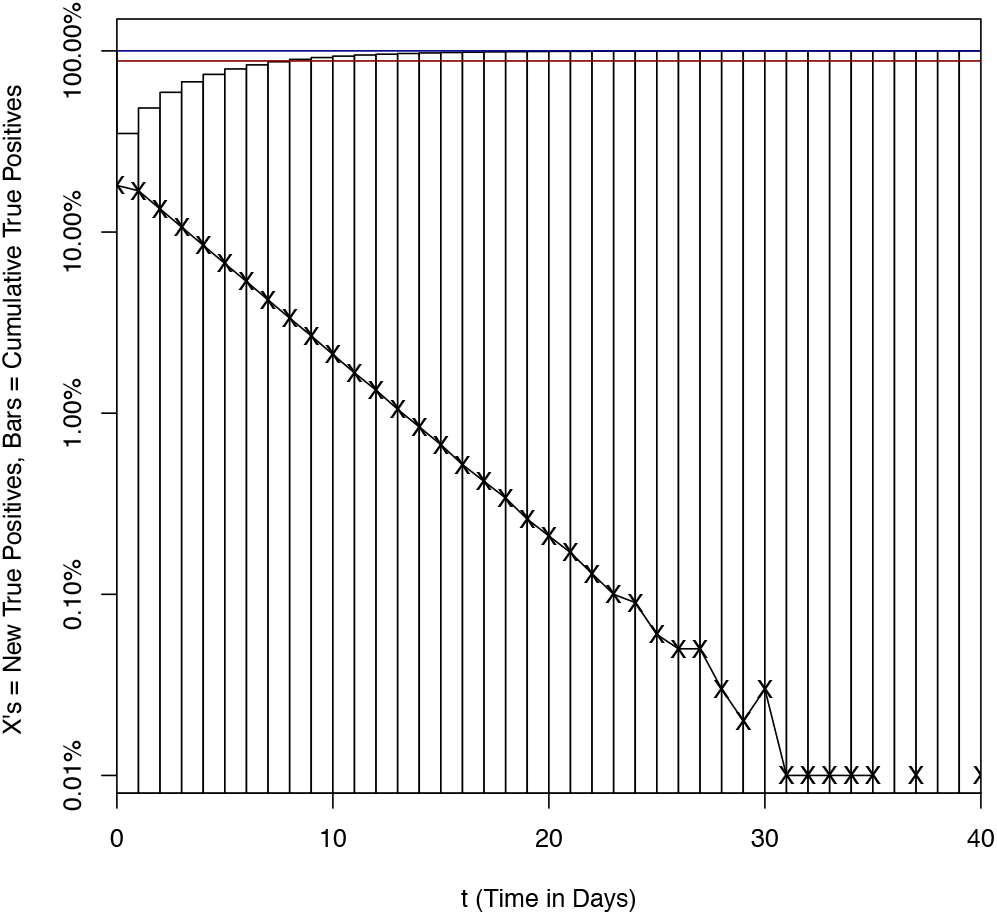
Exponential Decrease of New Infected Time Series

The three different types of new infected time series are shown above.

In each time series chart, the X’s connected with the black line represent the number of new infected each day t. The bars represent the cumulative infected. Of note is that once infected, in the time series members of the subpopulation are always counted as positive whether they remain infectious, or have recovered or died and are no longer infectious. The cumulative infected total represents the true total percentage of the tested population who are positive for the condition of ever having been infected.

Given a known true infected time series, using algebra and arithmetic the test results have been computed and presented in Table 1 as the ratio of Test Positive to True Positive. (Please see the Appendix for the calculations.) In the time series charts, these ratios are expressed as the red horizontal line, and compared to a “perfect” test in the blue horizontal lines. The charts y axis are logarithmic -- and understate the true significance of the undercount. The results clearly show that the ratio of test positive (test infected) to total infected is significantly reduced, in the exponential growth and the flat examples, compared to both (a) the actual number of infected using an accurate calculation; and (b) the naively computed number of infected using a single final sensitivity. This is because the Wadsworth Test, like most antibody tests, increases in sensitivity as the immune system responds to the infection over time. Therefore, if an analysis uses only an antibody test’s peak sensitivity (measured three or more weeks after infection), it would significantly underestimate the true infected rate. Further, there is a risk, particularly in test results that do not have their adjustments published, that public health officials, government executives, or the general public will assume naively that seroprevalence test results presented in summary form at press conferences represent an unbiased estimated of the population’s true cumulative infection percentage, when instead it is likely significantly higher. Therefore it is clear that care must be taken by the modeler in using test results, particularly those without published detail as to their calculation method or sensitivity and specificity.

**Table 1.** Time Series Characteristic vs Test Positivity

| Infected plus Recovered<br>Time Series<br>Characteristic | Test Positive /<br>True Positive<br>Ratio |
| --- | --- |
| Exponential Increase | 20.9% |
| Flat | 50.5% |
| Exponential Decrease | 88.0% |
| **Comparison to:** |  |
| Naive Use of Test Final<br>Sensitivity and Specificity<br>Instead of Test Curve | 89.0% |
| Naive Assumption of<br>Perfect Test | 100% |

### The SIRX Model

This paper will use a modified SIR model to demonstrate calibration. The model utilized for this example is a simplification of the SIR-X model of Maier and Brockmann [2], itself a generalization of the SIR model. In addition to the three basic SIR compartments of S (susceptible), I (infected), and R (recovered or deceased), there is an additional compartment X (confirmed and reported cases), which allows the model to connect to the confirmed cases reported by public health authorities.^3^

The model is defined by the following differential equations describing the flow between the compartments:

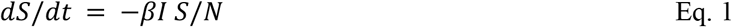

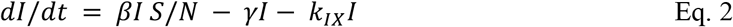

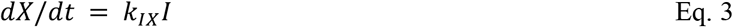

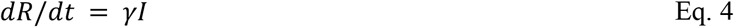

With the constraint:

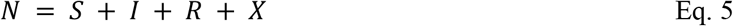

With definitions:

*S, I, R* ≡ Susceptible, Infected, or Recovered^4^ Population

*X* ≡ Confirmed Cases Population^5^ (assumed quarantined and thus unable to infect Susceptible Population)

*N* ≡ Total Population

*β* ≡ Transition rate from the Susceptible to the Infected compartment

γ ≡ Transition rate from the Infected to the Recovered compartment

*k_IX_* ≡ Transition rate from the Infected to Confirmed Cases compartment

*t* ≡ Time, in units of days for convenience

This model is a SIR model [3] with an extra compartment for the Confirmed cases and where the Confirmed cases are isolated until no longer infectious.

Because this paper’s purpose is to demonstrate calibration, for brevity the inherent weaknesses in the SIR model are immaterial and therefore ignored. Regardless, the calibration technique can be applied to more accurate and more complex models, including SEIR models, models that use gamma distributions for compartment distributions vs time, models that use or incorporate directly mortality, models that network individual infections, and so on. Examples include the Imperial College Model [4] and the IHME Model [5]. However, all models would benefit from more accurate calibration with seroprevalence data of which this simplified SIRX model is a mere example.

To simplify this example the γ transition factor has not been calibrated, but is simply assumed to be a constant of 1/8 representing an 8 day mean infectious period. 1/γ is equivalent to the mean time (in this case 8 days) in an exponential distribution for an infected individual to pass from the Infected compartment to the Recovered compartment (which included mortalities which are assumed to be no longer infectious). The 8 day mean time is both reasonable and sufficient to illustrate calibration to seroprevalence.

Within the SIRX model, as pointed out by Maier and Brockmann, the probability *Q_prob_* of a case ending up in the X Confirmed Cases compartment instead of the R compartment is:

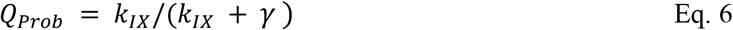

This is roughly equivalent but not the same as the ascertainment fraction, i.e. the ratio of confirmed cases to total infected plus recovered -- differing because of the delay of the infected from entering the Confirmed or Recovered compartments. Regardless, by adjusting *Q_Prob_* (or equivalently *k_IX_* through the linear relationship in Equation 6 above, the model may be calibrated to match seroprevalence as determined by survey.

### The Example Case Data

Daily Confirmed Case data was originally obtained from the New York City Department of Health by manually typing in the data in the daily reports [6], but that has been subsequently discontinued and does not make corrections for past reported data. NYC updated to a time series file [7] which has the advantage that past data are added as delayed reports are processed. The date used in the new NYC time series is the “date of diagnosis” which is presumably the same or slightly delayed from the date of a positive (PCR type) test being received by the medical practitioner. These data are then corroborated against data from the New York State Department of Health [8] by adding up the cases in the 5 counties comprising New York City (Kings, Queens, New York, Bronx, and Staten Island). Of note is that the NYS data’s date is “the date the test result was processed by the NYS Clinical Laboratory Reporting System”. All three case data sets are contained in the online code and data files for convenience. Visual observation shows that the NYS data are generally one day delayed from the NYC data; and that the NYC data have a larger weekly periodicity “noise” likely due to a relative lack of PCR testing or processing on Saturday and Sunday. The regression technique used in this paper largely averages out such noise, although a more sophisticated technique, based on either day of the week or tests on a day, could correct for the periodic weekly noise. The population for NYC was rounded to 8.623 mm from [9].

### The Computational Model

The model was built in the R Programming Language[10]. The SIRX model is encapsulated in a function sirX3() which when given initial conditions of *N*, *I_t_*_=_*_t_*_0_, *dI/dt* (at *t* = *t*0), *k_IX_*, *β*, and *γ*, will solve the differential equations (1) through (4) computationally for any number of days going forward using the Dormond-Prince Runge-Kutta 4th order method R method from the package deSolve as described in Soetaert et al [11]. This computational solution was checked by using a discrete numerical integration where each day is broken into 500 (arbitrarily small) timesteps (i.e. Euler’s method) as inspired by Anastassopoulou et al [12]).

The sirX3() function has an optional parameter which linearly changes the initial *β*_0_ by a multiplicative change factor *F_β_*_Δ_ (i.e. 0.5) over the fit interval from t_0_ to t_1_:

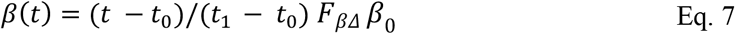

This is useful in fitting the data when a public health policy may have changed social distancing and self-isolation behavior as will be seen below.

### Fitting Case Data to Model

A relatively simplistic and (usually) stable method of fitting data is the least squares method. While there are essentially closed form solutions that produce an exact answer for linear models, non-linear models like SIRX cannot be solved using a linear regression. The problem is compounded when when *S/N* is significantly less than 1 or when a non-constant susceptible to infection transition rate (i.e. *β*) is assumed. (A changing *β* is equivalent to a changing effective reproduction number, *R_eff_* because *R_eff_* = *β/γ* in the SIR framework, and must be fit computationally by an algorithm).

Fortunately, Elzhov et al [13] has created within R an excellent Levenberg-Marquant least square fitting within the package minpack.lm called thru the nlsLM() function. The author has been iteratively improving the code algorithmically utilizing these functions for several weeks and has thus experienced and debugged multiple edge cases. In the interest of brevity, the code has been distilled down to a minimum example based around a calling function, runSIRX2, called from the command line.^6^ An example of running the code is contained in the function genFiguresAndTables() which will generate the figures and tables used in this paper.

In fitting the data, the algorithm fits the error of the model’s estimate against the logarithm of the total cases. This has some desirable computational properties: 1) same weighting of percentage error on large case count data points vs small case counts, i.e. 10,000 vs 500; 2) reducing to the common log-linear least square regression solution if *S/N* = 1. The author has shown (but it is beyond the scope of this paper) that a regression of

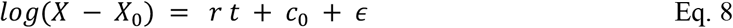

is the general “closed form” symbolic solution when *S/N* = 1 thereby giving the more complex SIRX non-linear least squares optimizer a computationally “nearby” starting set of parameters to bootstrap the regression. Where:

*r* ≡ the log linear regression’s coefficient on t, equivalent to the exponential growth rate

*c*_0_ ≡ the log linear regression’s constant

∈ ≡ the log linear regression’s residual error term

*X*_0_ ≡ the number of cases at t=0 in the regression.

If *X*_0_ is approximately known, for example in the initial growth phase of an epidemic as 1, Equation 8’s log-linear regression will produce a reasonable result. However, if *X*_0_ is unknown, for example when an epidemic’s new cases are declining exponentially, the regression cannot be formulated in a (log) linear form. In that event, to solve for the parameters *X*_0_, *A* and *r*, Equation 8 must be rearranged to a non-linear regression.^7^

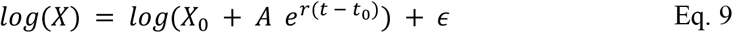

After this non-linear regression, with *X*_0_, *A*, and *r* estimated, the run on the SIRX model has good starting parameters and will normally converge to a solution.

The essential functional flow of the algorithm is:

1. Fetch the data;
2. Use simpler linear models (with *S/N* = 1 leaving purely exponential results) to guess initial parameters for the non-linear nlsLM model that will not result in a gradient singularity, thereby causing the model to fail;
3. As desired by the modeler, either use explicit inputs for *k_IX_* (equivalent to setting *Q_Prob_*) and the beta Change Factor to find the best fit for the current beta from the subset of the data.
4. If the modeler should so desire, the algorithm can find the best fit for *k_IX_* and/or the Beta Change Factor *F_βΔ_* in addition to *β* or *β*_0_ (the latter if using the Beta Change Factor *F_βΔ_*).

Of note is that the constants *β* and *k_IX_* are not constant in the real world over relatively long time periods. This is because social distancing behavior changes the interaction rate between the infected and the susceptible, and hence changes *β*; and because changes in screening behavior, contact tracing, or reticence to seek healthcare behavior also change with time. This changing of the *β* and *k_IX_* “constants” favors regression over shorter time periods. However, as was shown by the 3 examples based on a real-world seroprevalence test (like the Wadsworth test), a longer period is needed to take into account very significant changes in the test sensitivity. For the purposes of this calibration demonstration, this paper is essentially forced to use at least 30 days preceding the seroprevalence survey. A more accurate model could use more frequent seroprevalence sampling, and perhaps fatality data, to allow a piecewise approximation of the *β* and *k_IX_* regression parameters. This is beyond the scope of this paper.

An example of a calibration model run is in the Calibrating the SIRX to the Seroprevalence Survey section that will follow.

### The NYS Seroprevalence Data

The seroprevalence data for the example is taken from the Governor of New York, Andrew Cuomo’s, press conference prepared slides [14, 15, 16]. The final data used from this paper is taken from the slides (contained in the YouTubes in the references) which differ due to rounding from the Governor’s transcript remarks. The data is summarized below:

**Table 2.**
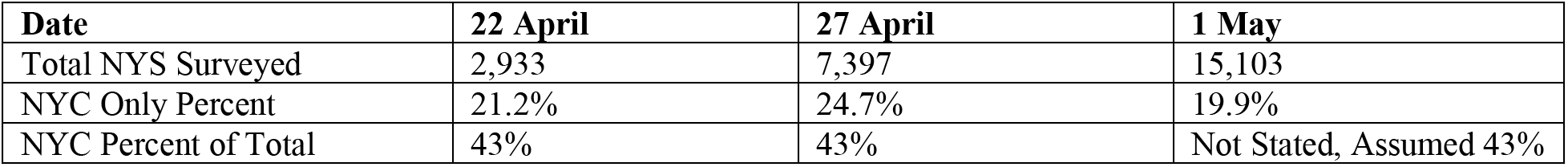
New York Antibody Survey Results

Of note is that the 22 April data, released on 23 April is stated as being “collected over two days”. It is assumed it was collected on 20-21 April. The 1 May data, released first on 2 May, is assumed collected 29-30 April. As it is unlikely that the NYC percentage positive test fraction could have declined by 4.8% (from 24.7% to 19.9%) in approximately doubling the sample size over 4 days, it is more likely that there was some unpublished adjustment to the serological survey data.^8^Further, the 2 May press conference indicated that the data was collected “over the past two weeks”.

Assuming a constant rate of collection, this paper will use the midpoint, 25 April, for calibration. However, a more accurate estimate could be obtained using the actual collections on each day for the calibration with some type of least squares fit to the data as will be described below. The survey administrator (i.e. a government public health authority), would have available any ex-post corrections made in the unpublished data. This author recommends that government and private survey collectors fully publish all data.

### The NYS “Wadsworth” Laboratory Antibody Seroprevalence Test

Governor Cuomo described the test as the New York State Department of Health “Wadsworth Center” independently developed antibody test [17]. The author’s research, prior to 1 May, did not find published data describing the specificity and accuracy of the Wadsworth Center’s test. However, the FDA has described test characteristics for a Wadsworth Center SARS-CoV-2 Antibody test in an Accelerated Emergency Use Authorization (“EUA”), which they released on 1 May 2020 [18, 19]. The tables below, reproduced directly from the EUA, describe the Wadsworth test sensitivity and specificity.^9^

**Table 3.** NYS Wadsworth Test Sensitivity Through “>20 Days from onset” Reproduced from “Table 2” in Reference [18]

| Days from onset | Samples tested (N) | RT-PCR result | NY SARS-CoV MIA Result as Compared to PCR |  |  | Sensitivity (95%CI) |
| --- | --- | --- | --- | --- | --- | --- |
|  |  |  | Positive | Indeter. | Negative |  |
| <7 | 179 | Pos | 32 | 30 | 117 | 32/179<br>17.9% (12.96% - 24.15%) |
| 7 – 10 | 67 | Pos | 21 | 19 | 27 | 21/67<br>31.3% (21.50% - 43.20%) |
| 11 – 15 | 47 | Pos | 23 | 4 | 20 | 23/47<br>48.9% (35.28% - 62.76%) |
| 16 – 20 | 126 | Pos | 62 | 19 | 45 | 62/126<br>49.2% (40.63% - 57.83%) |
| >20 | 334 | Pos | 265 | 28 | 41 | 265/334<br>79.3% (74.68% - 83.34%) |
| Total | 753 |  |  |  |  |  |

**Table 4.**
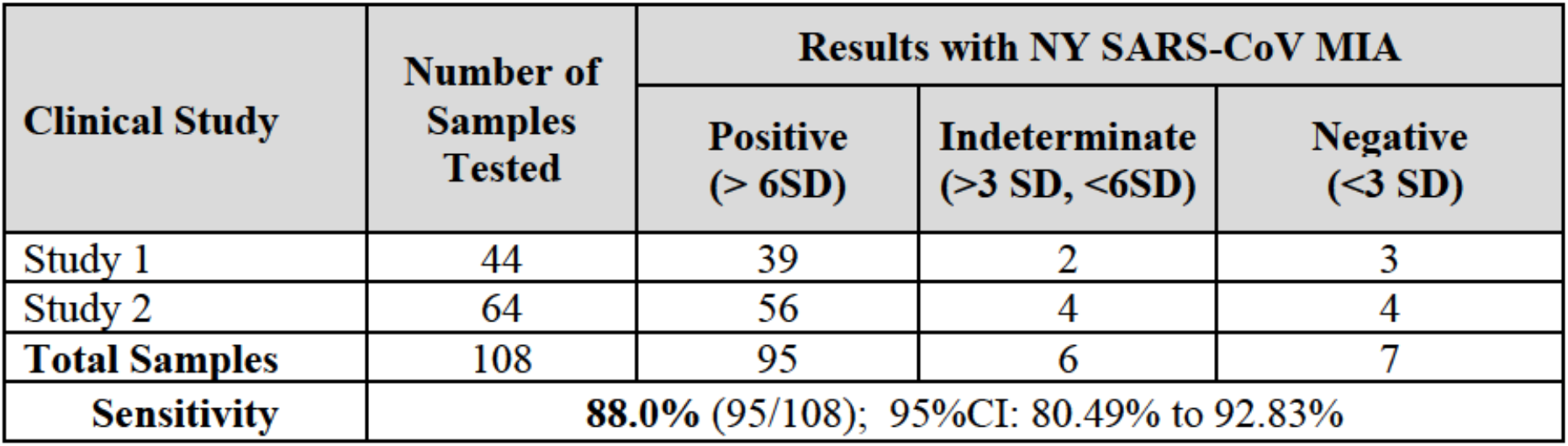
NYS Wadsworth Test Sensititivity “for samples collected at least 25 days after symptom onset” Reproduced from “Table 3” in Reference [18]

Note that in both NYS Wadsworth calibration tables above, the time units are days since symptom onset. For pmposes of this example, this paper will assume a 5 day incubation period between infection and symptom onset -- thus the sensitivity data need to be adjusted by adding 5 additional days to the left hand column. If the model used was a “SEIR” type model (compartments: **S**usceptible, **E**xposed, **I**nfectious, and **R**ecovered), where onset was being used as a proxy for infectious, this could be adjusted directly. Alternatively an additional compartment could be added if infectiousness begins before symptom onset but after infection, i.e. a SEIOR model (compartments: **S**usceptible, **E**xposed, **I**nfectious, **O**nset of symptoms, and **R**ecovered). Additionally, if the model has direct access to the testing results, additional granularity would be available.

The NYS Wadsworth EUA provided sensitivity curve is clearly non-linear between 11 and 20 days after onset. While this could be smoothed out with a curve fitting, if the model has direct access to the underlying data, a more accurate sensitivity curve could be constructed. Such accuracy and confidence interval information is beyond the time scope of this paper.

The specificity data (i.e. 100% less the false positive percentage) are given on page 7 of the EUA for a variety of sample sera and are reasonably assumed by the EUA to not vary with time since infection with SARS-Cov-2. In calibration, the total true positives for all 433 samples in the clinical specificity table (Table 5 of the EUA [18]) is used to calculate an average specificity for all 433 samples: 100% - 5/433 = 98.85%. Taking sensitivity and specificity together, these are used to make a “test calibration table” which is input into the R computational model:

**Table 5.** Wadsworth Calibration Test Sensitivity and Specificity

| **t start** | **t end** | **Sensitivity** | **Specificity** |
| --- | --- | --- | --- |
| 0 | 11.999... | 0.179 | 0.9885 |
| 12 | 15.999... | 0.313 | 0.9885 |
| 16 | 20.999... | 0.489 | 0.9885 |
| 21 | 25.999... | 0.492 | 0.9885 |
| 26 | 29.999... | 0.793 | 0.9885 |
| >=30 |  | 0.880 | 0.9885 |

### Test Calibration Sample Bias

As an additional caveat, please note that the Wadsworth Test clinical sensitivity tables are calibrated using 753 subjects from “several US clinical collection sites”, and using108 samples from Westchester County, collected at undisclosed times in March and April 2020. As the samples were taken from clinical PCR tests at a time when, due to rationing, PCR tests were generally limited to symptomatic individuals who sought out medical care presumably due to severity of illness, and where asymptomatic or mild cases would have difficulty in obtaining a PCR test; it is likely there is a calibration selection bias which increases the relative number of severe cases in the sample. Further, as the age distribution for those with severe disease is clearly biased older as seen in [20] than the general NYC population (seen in [9]), the Wadsworth Test calibration samples are likely biased older. If the model has direct access to the calibration population data, this bias can be eliminated.

### Using a SIRX run to Estimate a Seroprevalence Test Result

For a given SIRX model run, a table is generated of the true positives, that is the infected plus recovered in the general population that is being sampled in the NYS seroprevalence survey. For a given model run containing infected and recovered in the general population who have not been previously PCR (swab) tested positive, each such day has a certain number of newly infected on a given date in that population that can be calculated^10^ as:

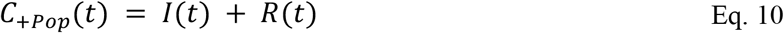

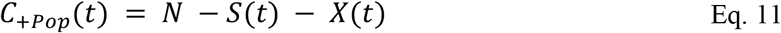

where the function at time notation *I*(*t*) indicates the value is a time series taken from the SIRX model and

C_+Pop_(*t*) ≡ the cumulative total infected at time *t* in the non “Confirmed-Case” population

Then, a careful application of Equation 33A from the Appendix^11^ yields the percentage that is tested positive and presumably reported by the Governor during the press conference. This is done by assuming that the *entire* non-Case Confirmed population is tested at the test date, so that

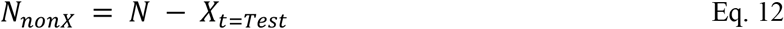

is used as the denominator in Equation 33A. While *X_t_*_=_*_Test_* is typically a small fraction of the total population *N*, it is somewhat significant. Therefore, Equation 33A becomes:

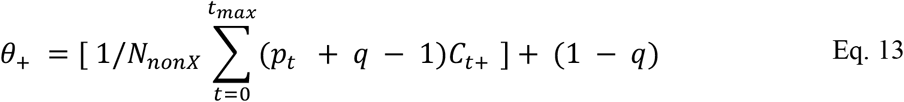

The *θ*_+_ is thus the fraction that would test positive if the survey encompassed the entire non-Confirmed Population and the test performed precisely as specified in Table 5. Of course, the actual test is a sample and the test is statistically described to have confidence intervals, so that statistical methods could be applied to derive a confidence interval on the test results. The fitting of the case data (using the least squares technique on a limited sample set) is itself subject to statistical estimation errors. But again, this paper is simply demonstrating the technique to derive a mean “point estimate” and the estimation of statistical error is beyond its (already lengthy) scope.

### Calibrating the SIRX to the Seroprevalence Survey

As a final step, the modeler flips a switch to get the runSIRX2() algorithm to calculate the seroprevalence; asking the model to adjust inputs for *k_IX_* (i.e. *Q_Prob_*) and the Beta Change Factor *F_βΔ_*; or alternatively asking the non-linear least squares algorithm to find a best fit to a seroprevalence that matches the survey data simultaneously with finding the best fit for the daily new cases *X* and the change in daily new cases *ΔX*. To do this in a single non-linear “regression” the weighting factor is increased for the seroprevalence target least squares error so that it is large enough to act as a constraint -- forcing the least squares algorithm to converge to a solution with seroprevalence very near (within 0.1%) of the survey result target. This yields a minimum least squares fit to the case data while simultaneously fitting the seroprevalence data, thereby calibrating the model. An example of this calibration is shown in Figure 4 below.

**Figure 4.**
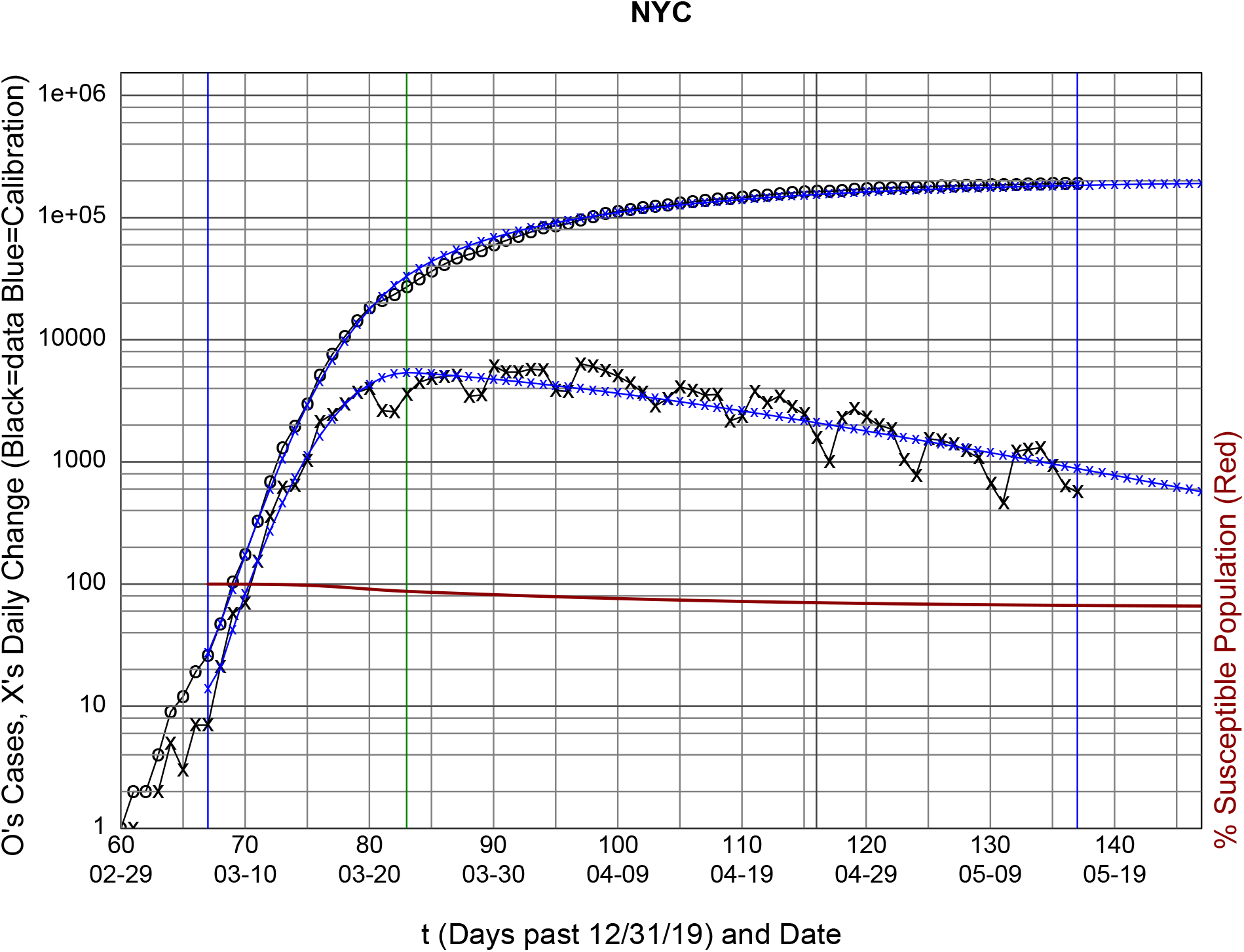
Calibrated to 19.9% Seroprevalence Survey SIRX using NYC Case Data

The black lines are the actual data with the upper black line (the O’s) being the reported cases in the data set (in this case NYC’s data from [7]), and the lower black line (the X’s) is the daily change in cases. The blue lines are the calibrated fit to the data. The red line is the *S/N*. i.e. the percentage still susceptible in the population reusing the 1 to 100 scale on the left Y axis.

The calibration needs additional inputs: 1) The mean “effective” date the seroprevalence data are collected (April 25th, the black vertical line), 2) the range of data for which to do the calibration (represented by the vertical blue lines from March 7th to May 16th) -- a section of time for which the data appear to be clean except for the weekly periodic (7 day) variation in tests^12^; and 3) the range of data for which *β*(*t*) is adjusted as per Equation 7 being also from March 7th and running until March 23rd (16 days = Δ*t_chg_* from the start of the calibration range). This *β*(*t*) range brackets the Governor’s “lockdown” social distancing order (March 20th). By manually varying Δ*t_chg_* in integer units, starting at the lockdown date, the minimum residual standard error may be found, so that the optimal Δ*t_chg_* is that date that has the minimum residual error. This may be done algorithmically, but is beyond the time scope of the code development associated with this paper.

Note that there may be timing differences of a few days between reporting of the PCR Swab Test data represented by the NYC times series data (the black lines) and the seroprevalence test dates which reported in the during the Governor’s press conferences. Also note that the change in *β*(*t*) is fit as if it is linear in the Δ*t_chg_* time interval and that this is an approximation to the hidden underlying process. Additional code could add a start date immediately before and after the lockdown date.

What can be seen is that 1) the SIRX model visually does a good job of fitting both the daily change in cases (lower blue line) and the cumulative daily cases (the upper blue line); and 2) the percent still susceptible is around 70% compared with the serological survey showing around 80%. The actual numbers can be seen in Table 6 below.

The model text output in Table 6 shows that a perfect seroprevalence test would show 28% positive^13^ in the total population on April 25th, and with 163,000 PCR reported infected (about 2%, presumed not in the seroprevalence survey) there are only about 71% remaining susceptible in the population. This continues to decrease slowly to around 67%.

**Table 6.** Model Run Data and Calibration

| | Date | t | C (Cases) | $\Delta C$ | I (Model) | X (Model) | $\Delta X$ (Model) | S% (Model) |
| --- | --- | --- | --- | --- | --- | --- | --- | --- |
| 1 | 2020-02-29 | 60 | 1 | NA | NA | NA | NA | NA |
| 2 | 2020-03-01 | 61 | 2 | 1 | NA | NA | NA | NA |
| 3 | 2020-03-02 | 62 | 2 | 0 | NA | NA | NA | NA |
| 4 | 2020-03-03 | 63 | 4 | 2 | NA | NA | NA | NA |
| 5 | 2020-03-04 | 64 | 9 | 5 | NA | NA | NA | NA |
| 6 | 2020-03-05 | 65 | 12 | 3 | NA | NA | NA | NA |
| 7 | 2020-03-06 | 66 | 19 | 7 | NA | NA | NA | NA |
| 8 | 2020-03-07 | 67 | 26 | 7 | 1556.4 | 27.2 | 13.9 | 99.98 |
| 9 | 2020-03-08 | 68 | 47 | 21 | 3275.0 | 47.9 | 20.7 | 99.95 |
| 10 | 2020-03-09 | 69 | 104 | 57 | 6567.3 | 90.2 | 42.3 | 99.91 |
| 11 | 2020-03-10 | 70 | 174 | 70 | 12547.5 | 172.9 | 82.6 | 99.82 |
| 12 | 2020-03-11 | 71 | 328 | 154 | 22834.3 | 326.6 | 153.7 | 99.68 |
| 13 | 2020-03-12 | 72 | 684 | 356 | 39563.7 | 598.9 | 272.3 | 99.44 |
| 14 | 2020-03-13 | 73 | 1304 | 620 | 65229.9 | 1058.2 | 459.3 | 99.06 |
| 15 | 2020-03-14 | 74 | 1947 | 643 | 102272.0 | 1795.3 | 737.1 | 98.50 |
| 16 | 2020-03-15 | 75 | 2980 | 1033 | 152387.4 | 2919.9 | 1124.6 | 97.72 |
| 17 | 2020-03-16 | 76 | 5103 | 2123 | 215676.0 | 4550.5 | 1630.6 | 96.71 |
| 18 | 2020-03-17 | 77 | 7557 | 2454 | 289882.2 | 6796.4 | 2245.9 | 95.45 |
| 19 | 2020-03-18 | 78 | 10534 | 2977 | 370090.3 | 9734.9 | 2938.5 | 94.01 |
| 20 | 2020-03-19 | 79 | 14241 | 3707 | 449146.0 | 13388.9 | 3654.0 | 92.46 |
| 21 | 2020-03-20 | 80 | 18251 | 4010 | 518806.8 | 17711.6 | 4322.8 | 90.90 |
| 22 | 2020-03-21 | 81 | 20886 | 2635 | 571301.3 | 22583.9 | 4872.2 | 89.44 |
| 23 | 2020-03-22 | 82 | 23461 | 2575 | 600808.1 | 27824.8 | 5240.9 | 88.18 |
| 24 | 2020-03-23 | 83 | 27019 | 3558 | 604447.0 | 33214.3 | 5389.5 | 87.20 |
| 25 | 2020-03-24 | 84 | 31514 | 4495 | 594742.4 | 38557.7 | 5343.4 | 86.38 |
| 26 | 2020-03-25 | 85 | 36348 | 4834 | 584540.7 | 43812.3 | 5254.6 | 85.59 |
| 27 | 2020-03-26 | 86 | 41371 | 5023 | 573901.2 | 48974.1 | 5161.8 | 84.81 |
| 28 | 2020-03-27 | 87 | 46463 | 5092 | 562870.7 | 54039.3 | 5065.2 | 84.05 |
| 29 | 2020-03-28 | 88 | 49919 | 3456 | 551495.4 | 59004.6 | 4965.3 | 83.32 |
| 30 | 2020-03-29 | 89 | 53442 | 3523 | 539820.6 | 63867.1 | 4862.6 | 82.61 |
| 31 | 2020-03-30 | 90 | 59543 | 6101 | 527890.6 | 68624.5 | 4757.4 | 81.92 |
| 32 | 2020-03-31 | 91 | 64971 | 5428 | 515748.4 | 73274.6 | 4650.1 | 81.25 |

<sup>13</sup> These figures in this paragraph are arbitrarily rounded to 1%. Also note that the precise calculation of a perfect seroprevalence test requires both the numerator and denominator be adjusted, i.e. $(\Sigma I - X) / (N - X)$ where $\Sigma I$ is the total cumulative infected. $S = S_{pct} N$ and $\Sigma I = N - S$ . For NYC $N \cong 8,623,000$ . In numbers carrying extra precision to make the arithmetic clear $(\Sigma I - X) / (N - X) \cong 2599527/8468394 \cong 28\%$ for April 25th.

Table 6 - Model Run Data and Calibration (Continued)
| | Date | t | C(Cases) | $\Delta C$ | I(Model) | X(Model) | $\Delta X(\text{Model})$ | S%(Model) |
| --- | --- | --- | --- | --- | --- | --- | --- | --- |
| 33 | 2020-04-01 | 92 | 70403 | 5432 | 503435.2 | 77815.7 | 4541.1 | 80.60 |
| 34 | 2020-04-02 | 93 | 76156 | 5753 | 490990.9 | 82246.4 | 4430.8 | 79.97 |
| 35 | 2020-04-03 | 94 | 81803 | 5647 | 478453.2 | 86565.8 | 4319.4 | 79.37 |
| 36 | 2020-04-04 | 95 | 85651 | 3848 | 465858.2 | 90773.2 | 4207.4 | 78.78 |
| 37 | 2020-04-05 | 96 | 89420 | 3769 | 453239.9 | 94868.3 | 4095.1 | 78.21 |
| 38 | 2020-04-06 | 97 | 95773 | 6353 | 440630.0 | 98850.9 | 3982.6 | 77.66 |
| 39 | 2020-04-07 | 98 | 101822 | 6049 | 428058.4 | 102721.3 | 3870.4 | 77.13 |
| 40 | 2020-04-08 | 99 | 107374 | 5552 | 415552.7 | 106480.0 | 3758.7 | 76.62 |
| 41 | 2020-04-09 | 100 | 112403 | 5029 | 403138.4 | 110127.6 | 3647.6 | 76.13 |
| 42 | 2020-04-10 | 101 | 116833 | 4430 | 390839.0 | 113665.1 | 3537.5 | 75.66 |
| 43 | 2020-04-11 | 102 | 120540 | 3707 | 378675.9 | 117093.5 | 3428.5 | 75.20 |
| 44 | 2020-04-12 | 103 | 123406 | 2866 | 366668.6 | 120414.3 | 3320.8 | 74.76 |
| 45 | 2020-04-13 | 104 | 126693 | 3287 | 354834.4 | 123628.8 | 3214.5 | 74.34 |
| 46 | 2020-04-14 | 105 | 130813 | 4120 | 343189.0 | 126738.7 | 3109.9 | 73.93 |
| 47 | 2020-04-15 | 106 | 134681 | 3868 | 331746.2 | 129745.7 | 3007.0 | 73.54 |
| 48 | 2020-04-16 | 107 | 138199 | 3518 | 320518.1 | 132651.7 | 2906.0 | 73.17 |
| 49 | 2020-04-17 | 108 | 141767 | 3568 | 309515.1 | 135458.7 | 2806.9 | 72.80 |
| 50 | 2020-04-18 | 109 | 143930 | 2163 | 298746.2 | 138168.6 | 2709.9 | 72.46 |
| 51 | 2020-04-19 | 110 | 146277 | 2347 | 288218.9 | 140783.7 | 2615.0 | 72.12 |
| 52 | 2020-04-20 | 111 | 150046 | 3769 | 277939.2 | 143306.0 | 2522.3 | 71.80 |
| 53 | 2020-04-21 | 112 | 153106 | 3060 | 267912.2 | 145737.9 | 2431.9 | 71.49 |
| 54 | 2020-04-22 | 113 | 156565 | 3459 | 258141.3 | 148081.5 | 2343.6 | 71.20 |
| 55 | 2020-04-23 | 114 | 159396 | 2831 | 248629.3 | 150339.2 | 2257.7 | 70.92 |
| 56 | 2020-04-24 | 115 | 161858 | 2462 | 239377.8 | 152513.3 | 2174.1 | 70.64 |
| 57 | 2020-04-25 | 116 | 163447 | 1589 | 230387.4 | 154606.2 | 2092.8 | 70.38 |
| 58 | 2020-04-26 | 117 | 164455 | 1008 | 221657.9 | 156620.1 | 2013.9 | 70.13 |
| 59 | 2020-04-27 | 118 | 166759 | 2304 | 213188.5 | 158557.4 | 1937.3 | 69.90 |
| 60 | 2020-04-28 | 119 | 169472 | 2713 | 204977.5 | 160420.3 | 1862.9 | 69.67 |
| 61 | 2020-04-29 | 120 | 171806 | 2334 | 197022.8 | 162211.2 | 1790.9 | 69.45 |
| 62 | 2020-04-30 | 121 | 173809 | 2003 | 189321.5 | 163932.4 | 1721.2 | 69.24 |
| 63 | 2020-05-01 | 122 | 175674 | 1865 | 181870.3 | 165586.1 | 1653.7 | 69.03 |
| 64 | 2020-05-02 | 123 | 176722 | 1048 | 174665.7 | 167174.4 | 1588.4 | 68.84 |
| 65 | 2020-05-03 | 124 | 177499 | 777 | 167703.4 | 168699.7 | 1525.2 | 68.66 |
| 66 | 2020-05-04 | 125 | 179043 | 1544 | 160979.1 | 170164.0 | 1464.3 | 68.48 |
| 67 | 2020-05-05 | 126 | 180552 | 1509 | 154488.1 | 171569.3 | 1405.4 | 68.31 |
| 68 | 2020-05-06 | 127 | 181951 | 1399 | 148225.4 | 172917.9 | 1348.6 | 68.15 |
| 69 | 2020-05-07 | 128 | 183191 | 1240 | 142186.0 | 174211.7 | 1293.8 | 67.99 |
| 70 | 2020-05-08 | 129 | 184276 | 1085 | 136364.5 | 175452.6 | 1240.9 | 67.84 |
| 71 | 2020-05-09 | 130 | 184944 | 668 | 130755.6 | 176642.6 | 1190.0 | 67.70 |
| 72 | 2020-05-10 | 131 | 185403 | 459 | 125353.7 | 177783.5 | 1140.9 | 67.56 |
| 73 | 2020-05-11 | 132 | 186622 | 1219 | 120153.2 | 178877.3 | 1093.7 | 67.43 |
| 74 | 2020-05-12 | 133 | 187885 | 1263 | 115148.6 | 179925.5 | 1048.2 | 67.31 |
| 75 | 2020-05-13 | 134 | 189189 | 1304 | 110334.2 | 180930.0 | 1004.5 | 67.19 |
| 76 | 2020-05-14 | 135 | 190123 | 934 | 105704.4 | 181892.4 | 962.4 | 67.08 |
| 77 | 2020-05-15 | 136 | 190761 | 638 | 101253.5 | 182814.4 | 922.0 | 66.97 |
| 78 | 2020-05-16 | 137 | 191327 | 566 | 96976.0 | 183697.5 | 883.1 | 66.86 |
| 79 | 2020-05-17 | 138 | NA | NA | 92866.4 | 184543.2 | 845.7 | 66.76 |
| 80 | 2020-05-18 | 139 | NA | NA | 88919.2 | 185353.0 | 809.8 | 66.67 |
| 81 | 2020-05-19 | 140 | NA | NA | 85129.0 | 186128.4 | 775.4 | 66.58 |
| 82 | 2020-05-20 | 141 | NA | NA | 81490.5 | 186870.7 | 742.3 | 66.49 |
| 83 | 2020-05-21 | 142 | NA | NA | 77998.4 | 187581.2 | 710.5 | 66.41 |
| 84 | 2020-05-22 | 143 | NA | NA | 74647.8 | 188261.2 | 680.0 | 66.33 |
| 85 | 2020-05-23 | 144 | NA | NA | 71433.6 | 188911.9 | 650.8 | 66.25 |
| 86 | 2020-05-24 | 145 | NA | NA | 68350.8 | 189534.6 | 622.7 | 66.18 |
| 87 | 2020-05-25 | 146 | NA | NA | 65394.8 | 190130.5 | 595.8 | 66.11 |
| 88 | 2020-05-26 | 147 | NA | NA | 62738.0 | 190717.6 | 569.2 | 65.90 |

**Run 1.**
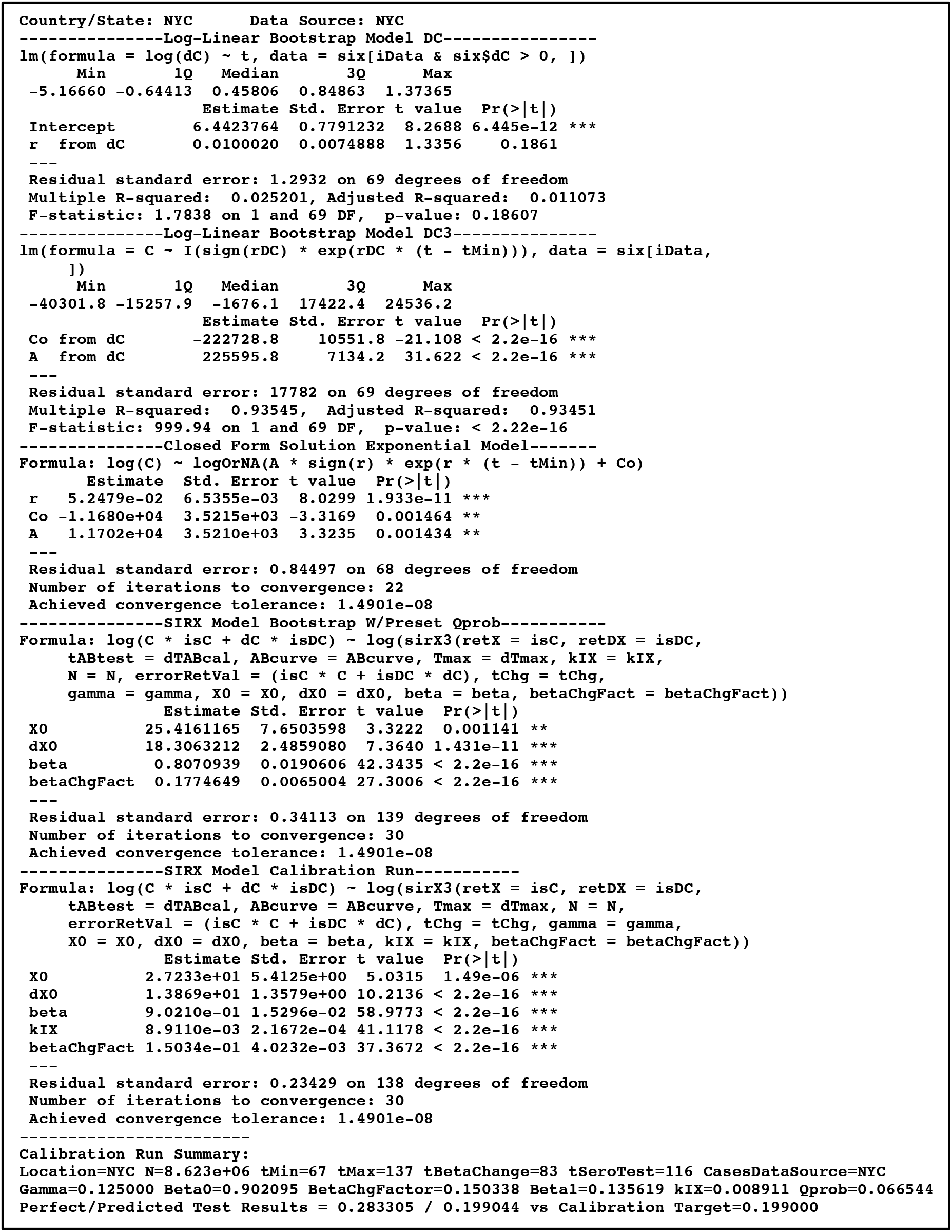
R Model Run Output^14^

## Discussion and Conclusion

Why is the model’s estimate of true infected plus recovered 40% higher than in the reported data?

The difference is due to a calibration with the FDA’s published sensitivity table from the NYS Wadsworth Lab test against a modelled slowly growing total number of infected plus recovered in the general population. While it is not known if this seroprevalence data were already calibrated (i.e. adjusted to present the population true infected plus recovered percentage) when presented by the New York Governor, it is important for modelers to ask the question. It is important to ask for full disclosure of the method of calibrating the test. If the test was not calibrated, a large underestimation of the seroprevalence would result, and hence getting to and achieving so called herd immunity would appear to be a more difficult task that must be more slowly approached.

Further, this implies that the infection fatality ratio (the IFR) may be overestimated by a similar factor of around 40% if the calibration is accurate. It is important for our public health leadership to have such information to make accurate estimates of the effects of various strategies. As other models use IFR as an input together with mortality data to estimate the current state and future evolution of the epidemic, it is important to estimate more accurately these data, suitably adjusted for delayed sensitivity, so as to draw more accurate conclusions from those models.

This paper does not purport to be an accurate estimate of seroprevalence, true cumulative infected or IFR. It serves to technically assist other modelers who may incorporate its techniques to understand the epidemiological situation. This paper’s seroprevalence calibration technique, especially after refinement, checking for inaccuracies, and integration into more advanced models, is applicable in cities, smaller states, and smaller countries where seroprevalence data is available, and where the epidemiological parameters like social interaction are fairly uniform. Epidemiologically heterogenous geographic units would necessarily need to be subdivided into more homogenous subunits for analysis.

As of the date of this writing, in the opinion of the author, it is of utmost urgency to continue seroprevalence survey collection and to use it for model calibration, so that we can better understand, collectively, where we are now, and where the epidemic will likely take us.

### Weaknesses

1. Instability of *k_IX_* and *β* over time.
2. There is bias in the sample used to produce the seropositivity test compared to the survey population.
3. Too many degrees of freedom in the regression may require additional seroprevalence data points, or incorporation of mortality data to determine both *k_IX_* and *β*.
4. Inaccurate estimation of sensitivity and specificity increases error.
5. Non-uniform subpopulations would need to be subdivided into multiple uniform analyses.
6. Noise in the case data, for example from different rates of testing, needs to be mitigated further
7. Lack of confidence intervals would allow naive application of this model to draw significantly inaccurate conclusions.
8. This paper was written by one person and published without peer review -- there may be significant errors.
9. ***This paper should not be used directly to support any epidemiological conclusion without professional review. It is designed to be an element that can be incorporated into other professional models.***

## Data Availability

All data is available both on Github an in the supplemental files.

https://github.com/becare-rocket/calibratingSIRX

## Math Appendix: Calculating Test Results from the 3 S’s: Seropositivity, Sensitivity and Specificity

Let

*N* ≡ Total number tested

*x* ≡ Number of tested positives that are condition positive (true positives)

*y* ≡ Number of tested negatives that are condition negative (true negatives)

*x* ≡ Number of tested positive that are condition negative (false positives)

*y’* ≡ Number of tested negative that are condition positive (false negatives)

*p* ≡ Sensitivity of the test

*q* ≡ Specificity of the test

*T*_+_ ≡ Total number tested positive (regardless of the true condition)

*T*_−_ ≡ Total number tested negative (regardless of the true condition)

*S*_+_ ≡ Fraction of tested who are (true condition) seropositive

*C*_+_ ≡ number of tested who are condition positive (regardless of the test result)

*C_−_* = number of tested who are condition negative (regardless of the test result)

By definition:

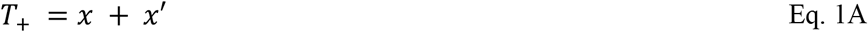

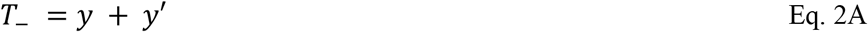

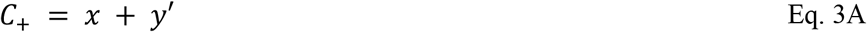

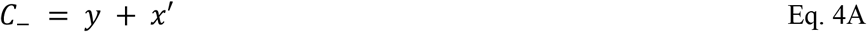

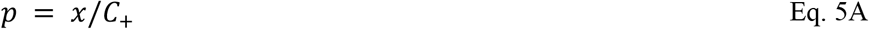

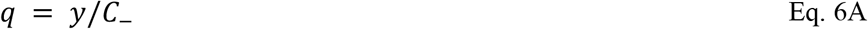

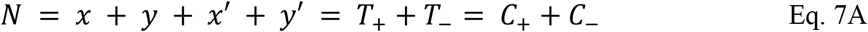

Combining the definitions in Eq. 3A and 4A with 5A and 6A:

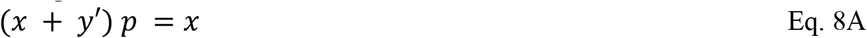

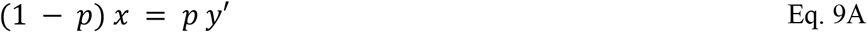

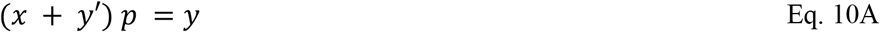

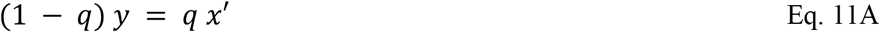

Combining Eq. 2A and 9A; and 1A and 11A respectively:

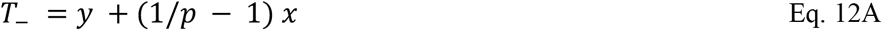

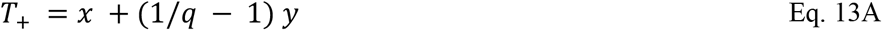

Rearranging Eq. 12A:

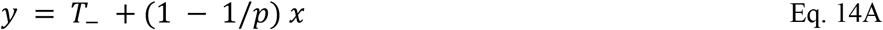

Substituting Eq. 14A into 13A and then expanding, simplifying, rearranging and solving for x:

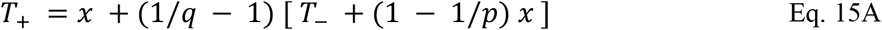

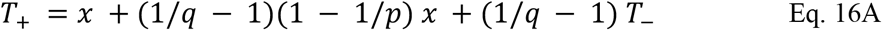

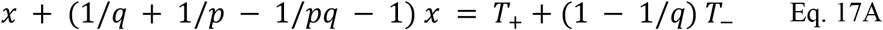

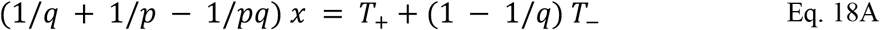

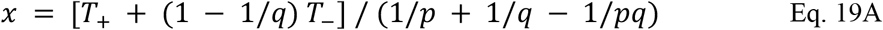

By symmetry:

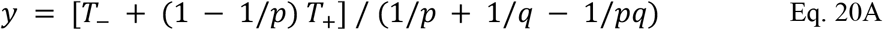

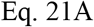

By rearranging Eq. 9A to solve for y’ and substituting the definition of *C_+_* from Eq. 3A, and then simplifying:

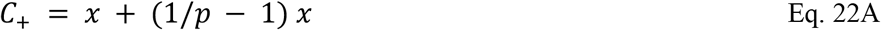

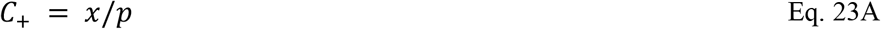

Substituting Eq. 19A into 23A and multiplying through by 1/*p*:

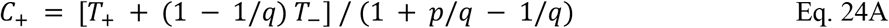

Noting the definition of *N = T+ + T_* and rearrange the definition to substitute for T+ in Eq, 24A:

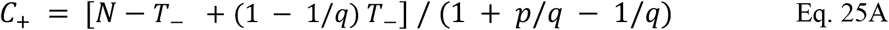

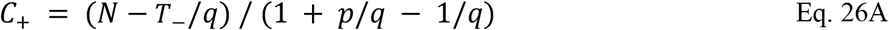

Using again the definition *N =* T_+_ + T_−_ and substituting for *T*_−_:

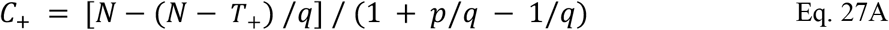

Simplifying and then solving for *T+*

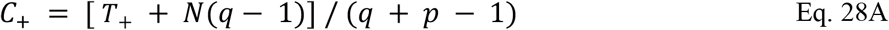

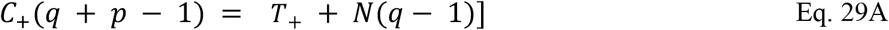

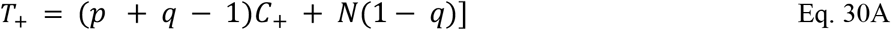

Thus, if one knows with certainty the specificity and sensitivity for any test population and the number who have been tested and are actually positive, the number that are tested positive are known.^15^

This can be used to illustrate the difference between naively taking the positive antibody percentage of a test as an estimate of the true positive antibody percentage of the same test population. It can also be compared to the assumption of a single sensitivity against an actual calibrated test that has increasing sensitivity dependent on the number of days since infection.

Note that if the sum of the sensitivity and specificity is less than 1 (i.e. their sum is less than 100%), the antibody test is unusable (i.e. bad) as the coefficient on *C*_+_ is negative, so that a higher condition positive result would impossibly create a lower test positive result. This corresponds to a receiver operating characteristic (ROC) point in the lower right hand half of the plot that is worse than a random guess [21], i.e. more wrong than right.

Also note *if the specificity is a perfect 100%* Eq. 30A reduces to simplified forms:

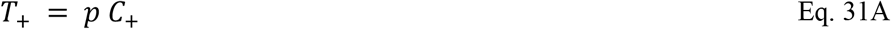

Equation 31A is occasionally useful, but for example in the cases where specificity is even just 5% below 100% (i.e. 95%), this simplified equation yields significant errors.

A simple example will show how this information can be used to calculate the test results from known seropositivity:

Assumption: Specificity is known precisely as 0.99 and does not vary with time. Sensitivity is exactly according to this table of the time post infection:

**Table 7A.** Simplified (Rounded) Sensitivity Table

| Days Since Infection<br>Interval Start | Days Since Infection<br>Interval End | Sensitivity |
| --- | --- | --- |
| 0 | 11.999... | 0.20 |
| 12 | 20.999... | 0.30 |
| 21 | 25.999... | 0.50 |
| 26 | 29.999... | 0.80 |
| 30 | No end | 0.90 |

This is a rounded to the nearest 10% version of the NY Wadsworth Test (see this paper’s Table 5 - Wadsworth Calibration Test Sensitivity and Specificity). When the tested subjects consists of a pool with different infection dates, Equation 30A cannot be used to compute the tested positive for the test in aggregate. Instead, each “vintage” of infection must be used to individually calculate the tested positive for that vintage, with the total of tested positive for each vintage then summed up to get the total tested positive for the survey. Additionally, the false positives must be included for the cohort of the test that has never been infected. Let

*θ*_+_ ≡ The percentage of the entire test population that is test positive (of all vintages)

*S*_+_ ≡ The percentage of the entire test population that is condition (truly) positive (of all vintages)

By definition

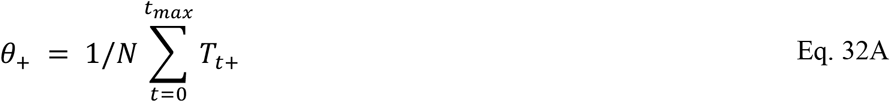

Or by substitution of Eq. 30A into Eq. 32, and then expanding and simplifying:

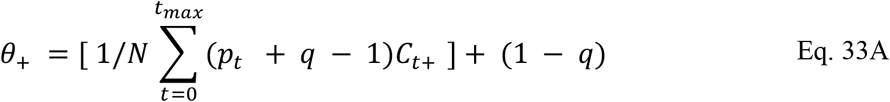

The formula in 33A is easy to apply. 1) Use a model to estimate the true infected at each time (in days) *C*_+_. i.e. at the 25 day vintage say there are 4 infected. 2) Multiply that by the coefficient which is the sum of the sensitivity *p_t_* for that 25 day vintage, plus the specificity, less 1. 3) Sum that up for all vintages. Note that for those beyond the maximum (in the table above, 30 or more days), they can be included all together. 4) Divide that sum by the total number of tested (regardless of test result or true infected or not infected state. 5) Add to the result 1 minus the specificity (which is assumed to be the same at all vintages. The result is the percentage of the entire test population that is expected to test positive assuming the specificity and sensitivity is exactly correct.

In the following examples, it will be convenient computationally to split *T*_+_ in Eq. 30A into a left and right half, i.e.

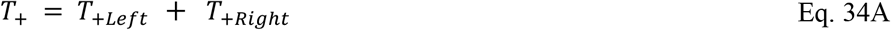

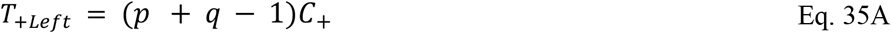

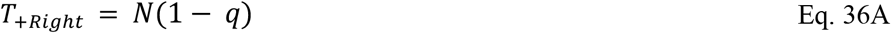

By inspection one can see that *T_+Left_* is the component of the positive tests due to actual condition positive, and that *T_+Right_* is the component of the positive tests due to (imperfect) specificity without regard for condition.

Similarly, it is convenient to split the percentage test results in Equation 33A into left and right component:

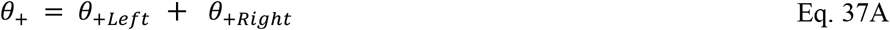

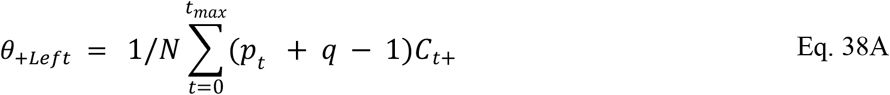

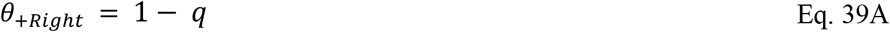

An example is below for a cumulative infected population doubling every 3 days arbitrarily cutoff at 10000 total infected, tested at 40 days since Day 0:

**Table 8.** Infections Doubling Every 3 Days Example

| $t$ | $\Delta t$ | $p_t$ | $\sum_{t=0}^t C_{t+}$ | $C_{t+}$ | $T_{+Left}$ |
| --- | --- | --- | --- | --- | --- |
| 0 | 40 | 0.9 | 1 | 1 | 0.89 |
| 1 | 39 | 0.9 | 1 | 0 | 0.00 |
| 2 | 38 | 0.9 | 1 | 0 | 0.00 |
| 3 | 37 | 0.9 | 2 | 1 | 0.89 |
| 4 | 36 | 0.9 | 2 | 0 | 0.00 |
| 5 | 35 | 0.9 | 3 | 1 | 0.89 |
| 6 | 34 | 0.9 | 4 | 1 | 0.89 |
| 7 | 33 | 0.9 | 5 | 1 | 0.89 |
| 8 | 32 | 0.9 | 6 | 1 | 0.89 |
| 9 | 31 | 0.9 | 8 | 2 | 1.78 |
| 10 | 30 | 0.9 | 10 | 2 | 1.78 |
| 11 | 29 | 0.8 | 12 | 2 | 1.58 |
| 12 | 28 | 0.8 | 16 | 4 | 3.16 |
| 13 | 27 | 0.8 | 20 | 4 | 3.16 |
| 14 | 26 | 0.8 | 25 | 5 | 3.95 |
| 15 | 25 | 0.5 | 32 | 7 | 3.43 |
| 16 | 24 | 0.5 | 40 | 8 | 3.92 |
| 17 | 23 | 0.5 | 50 | 10 | 4.90 |
| 18 | 22 | 0.5 | 64 | 14 | 6.86 |
| 19 | 21 | 0.5 | 80 | 16 | 7.84 |
| 20 | 20 | 0.3 | 101 | 21 | 6.09 |
| 21 | 19 | 0.3 | 128 | 27 | 7.83 |
| 22 | 18 | 0.3 | 161 | 33 | 9.57 |
| 23 | 17 | 0.3 | 203 | 42 | 12.18 |
| 24 | 16 | 0.3 | 256 | 53 | 15.37 |
| 25 | 15 | 0.3 | 322 | 66 | 19.14 |
| 26 | 14 | 0.3 | 406 | 84 | 24.36 |
| 27 | 13 | 0.3 | 512 | 106 | 30.74 |
| 28 | 12 | 0.3 | 645 | 133 | 38.57 |
| 29 | 11 | 0.2 | 812 | 167 | 31.73 |
| 30 | 10 | 0.2 | 1024 | 212 | 40.28 |
| 31 | 9 | 0.2 | 1290 | 266 | 50.54 |
| 32 | 8 | 0.2 | 1625 | 335 | 63.65 |
| 33 | 7 | 0.2 | 2048 | 423 | 80.37 |
| 34 | 6 | 0.2 | 2580 | 532 | 101.08 |
| 35 | 5 | 0.2 | 3250 | 670 | 127.30 |
| 36 | 4 | 0.2 | 4096 | 846 | 160.74 |
| 37 | 3 | 0.2 | 5160 | 1064 | 202.16 |
| 38 | 2 | 0.2 | 6501 | 1341 | 254.79 |
| 39 | 1 | 0.2 | 8191 | 1690 | 321.10 |
| 40 | 0 | 0.2 | 10000 | 1809 | 343.71 |
| Subtotal |  |  |  | 10000 | 1989.00 |
| $\theta_{+Left}$ | | | | | 19.9% |
| $\theta_{+Right}$ | | | | | 1.0% |
| $\theta_{+}$ | | | | | 20.9% |

As can be noted, the calculated total test positives *θ*_+_ is only 20.89%, whereas the total true positives is 100%. This ratio, of approximate 5x the true positives to test positives is exaggerated because 1) the number of infections is rapidly growing; and 2) in the end 100% of this hypothetical test population is infected.

**Table 9.** Flat New Cases Example

| $t$ | $\Delta t$ | $p_t$ | $\sum_{t=0}^t C_{t+}$ | $C_{t+}$ | $T_{+Left}$ |
| --- | --- | --- | --- | --- | --- |
| 0 | 40 | 0.9 | 244 | 244 | 217.16 |
| 1 | 39 | 0.9 | 488 | 244 | 217.16 |
| 2 | 38 | 0.9 | 732 | 244 | 217.16 |
| 3 | 37 | 0.9 | 976 | 244 | 217.16 |
| 4 | 36 | 0.9 | 1220 | 244 | 217.16 |
| 5 | 35 | 0.9 | 1464 | 244 | 217.16 |
| 6 | 34 | 0.9 | 1708 | 244 | 217.16 |
| 7 | 33 | 0.9 | 1952 | 244 | 217.16 |
| 8 | 32 | 0.9 | 2196 | 244 | 217.16 |
| 9 | 31 | 0.9 | 2440 | 244 | 217.16 |
| 10 | 30 | 0.9 | 2684 | 244 | 217.16 |
| 11 | 29 | 0.8 | 2928 | 244 | 192.76 |
| 12 | 28 | 0.8 | 3172 | 244 | 192.76 |
| 13 | 27 | 0.8 | 3416 | 244 | 192.76 |
| 14 | 26 | 0.8 | 3660 | 244 | 192.76 |
| 15 | 25 | 0.5 | 3904 | 244 | 119.56 |
| 16 | 24 | 0.5 | 4148 | 244 | 119.56 |
| 17 | 23 | 0.5 | 4392 | 244 | 119.56 |
| 18 | 22 | 0.5 | 4636 | 244 | 119.56 |
| 19 | 21 | 0.5 | 4880 | 244 | 119.56 |
| 20 | 20 | 0.3 | 5124 | 244 | 70.76 |
| 21 | 19 | 0.3 | 5368 | 244 | 70.76 |
| 22 | 18 | 0.3 | 5612 | 244 | 70.76 |
| 23 | 17 | 0.3 | 5856 | 244 | 70.76 |
| 24 | 16 | 0.3 | 6100 | 244 | 70.76 |
| 25 | 15 | 0.3 | 6344 | 244 | 70.76 |
| 26 | 14 | 0.3 | 6588 | 244 | 70.76 |
| 27 | 13 | 0.3 | 6832 | 244 | 70.76 |
| 28 | 12 | 0.3 | 7076 | 244 | 70.76 |
| 29 | 11 | 0.2 | 7320 | 244 | 46.36 |
| 30 | 10 | 0.2 | 7564 | 244 | 46.36 |
| 31 | 9 | 0.2 | 7808 | 244 | 46.36 |
| 32 | 8 | 0.2 | 8052 | 244 | 46.36 |
| 33 | 7 | 0.2 | 8296 | 244 | 46.36 |
| 34 | 6 | 0.2 | 8540 | 244 | 46.36 |
| 35 | 5 | 0.2 | 8784 | 244 | 46.36 |
| 36 | 4 | 0.2 | 9028 | 244 | 46.36 |
| 37 | 3 | 0.2 | 9272 | 244 | 46.36 |
| 38 | 2 | 0.2 | 9516 | 244 | 46.36 |
| 39 | 1 | 0.2 | 9760 | 244 | 46.36 |
| 40 | 0 | 0.2 | 10000 | 240 | 45.60 |
| Subtotal |  |  |  | 10000 | 4950.00 |
| $\theta_{+Left}$ | | | | | 49.5% |
| $\theta_{+Right}$ | | | | | 1.0% |
| $\theta_{+}$ | | | | | 50.5% |

In this example, daily new infections are constant with the exception of the final day, where they are reduced from 244 to 240 to make the example equal exactly 10,000 for the period. Note that the percentage infected detected by the test *θ*_+_ is half the true infected rate.

**Table 10.** Infections Decreasing at 3 days Half-Life Example

| $t$ | $\Delta t$ | $p_t$ | $\sum_{t=0}^t C_{t+}$ | $C_{t+}$ | $T_{+Left}$ |
| --- | --- | --- | --- | --- | --- |
| 0 | 40 | 0.9 | 1809 | 1809 | 1610.01 |
| 1 | 39 | 0.9 | 3499 | 1690 | 1504.10 |
| 2 | 38 | 0.9 | 4840 | 1341 | 1193.49 |
| 3 | 37 | 0.9 | 5904 | 1064 | 946.96 |
| 4 | 36 | 0.9 | 6750 | 846 | 752.94 |
| 5 | 35 | 0.9 | 7420 | 670 | 596.30 |
| 6 | 34 | 0.9 | 7952 | 532 | 473.48 |
| 7 | 33 | 0.9 | 8375 | 423 | 376.47 |
| 8 | 32 | 0.9 | 8710 | 335 | 298.15 |
| 9 | 31 | 0.9 | 8976 | 266 | 236.74 |
| 10 | 30 | 0.9 | 9188 | 212 | 188.68 |
| 11 | 29 | 0.8 | 9355 | 167 | 131.93 |
| 12 | 28 | 0.8 | 9488 | 133 | 105.07 |
| 13 | 27 | 0.8 | 9594 | 106 | 83.74 |
| 14 | 26 | 0.8 | 9678 | 84 | 66.36 |
| 15 | 25 | 0.5 | 9744 | 66 | 32.34 |
| 16 | 24 | 0.5 | 9797 | 53 | 25.97 |
| 17 | 23 | 0.5 | 9839 | 42 | 20.58 |
| 18 | 22 | 0.5 | 9872 | 33 | 16.17 |
| 19 | 21 | 0.5 | 9899 | 27 | 13.23 |
| 20 | 20 | 0.3 | 9920 | 21 | 6.09 |
| 21 | 19 | 0.3 | 9936 | 16 | 4.64 |
| 22 | 18 | 0.3 | 9950 | 14 | 4.06 |
| 23 | 17 | 0.3 | 9960 | 10 | 2.90 |
| 24 | 16 | 0.3 | 9968 | 8 | 2.32 |
| 25 | 15 | 0.3 | 9975 | 7 | 2.03 |
| 26 | 14 | 0.3 | 9980 | 5 | 1.45 |
| 27 | 13 | 0.3 | 9984 | 4 | 1.16 |
| 28 | 12 | 0.3 | 9988 | 4 | 1.16 |
| 29 | 11 | 0.2 | 9990 | 2 | 0.38 |
| 30 | 10 | 0.2 | 9992 | 2 | 0.38 |
| 31 | 9 | 0.2 | 9994 | 2 | 0.38 |
| 32 | 8 | 0.2 | 9995 | 1 | 0.19 |
| 33 | 7 | 0.2 | 9996 | 1 | 0.19 |
| 34 | 6 | 0.2 | 9997 | 1 | 0.19 |
| 35 | 5 | 0.2 | 9998 | 1 | 0.19 |
| 36 | 4 | 0.2 | 9998 | 0 | 0.00 |
| 37 | 3 | 0.2 | 9999 | 1 | 0.19 |
| 38 | 2 | 0.2 | 9999 | 0 | 0.00 |
| 39 | 1 | 0.2 | 9999 | 0 | 0.00 |
| 40 | 0 | 0.2 | 10000 | 1 | 0.19 |
|  |  |  | Subtotal | 10000 | 8700.80 |
| | | | $\theta_{+Left}$ | | 87.0% |
| | | | $\theta_{+Right}$ | | 1.0% |
| | | | $\theta_{+}$ | | 88.0% |

Even in an example where infections are decreasing at an exponential rate, with a 100% true infection rate, the antibody test only gets an 88% total tested positives. However, the amount of underestimation is only 100-88=12% in this case, vs. 100-21=79% in the growing case or 100-50=50%% in the flat case.

This stark difference in underestimation of the true infected percentage depending on the growth or decline of the true infected population time series demonstrates that calibration requires an estimation of the rate of growth or the decline in growth of the true infected time series.

## Footnotes

3 Note that Maier and Brockmann provide for a direct transition between the S and the R compartment to explain subexponential growth. This S to R transition is not used in this paper’s model.

4 Note that the Recovered Population includes those who have died. This population had been infected but is no longer infectious.

5 Similarly the (Confirmed) Case Population includes those who had been confirmed as positive but subsequently died or have recovered, so that they are no longer infectious to the Susceptible population.

6 An example of running the code is contained in the function genFiguresAndTables() which will generate the figures and tables used in this paper.

7 The reader may note that *t*_0_ is not known. It is set by convenience as the start of the data’s interval to be analyzed, noting that the regression solved non-linear regression *A*, and *r* constants can compensate for any choice of *t*_0_ because 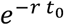 is also a constant.

8 27 April data: 7,397 × 24.7% × 43%= 786 positive. 1 May data: 15,103 × 19.9% × 43% = 1,292 positive. Therefore, during the 4 day period from 27 April to 1 May, there would only be 1,292 - 786 = 506 new positives out of 43% × (15,103 - 7,397) = 3,314 new tests or 506/3,314=15.3% positives, a decline of 24.7-15.3=9.4% in six days. This is statistically unlikely in such a sample size if the population and test were uniform, and therefore indicates a likely undisclosed adjustment to the data. This paper will use the 19.9% midpoint number is throughout.

9 Note that there is a typographical error in the EUA which refers to Table 3 as Table 4..

10 This estimate may be slightly inaccurate as some of the *X*(*t*) may have a different time series distribution vs. the *I*(*t*) + *R*(*t*) distribution -- because *R*(*t*) and *X*(*t*) are delayed compared to *I*(*t*). This in turn is due to the fact that the SIRX population must flow thru *I* to get to *R* and *X* compartments. However, to the extent that the ratio of *X*(*t*)/[*I*(*t*) + *R*(*t*)] is small and the percentage variation between Δ*X*(*t*)/*X*(*t*) is essentially by definition similar to Δ*R*(*t*)/*R*(*t*) (because Δ*X*(*t*) is proportion to *I*(*t*), as is Δ*R*(*t*)), this effect is likely to be small. Calculation of this value is beyond the time scope of this paper.

11 Please see the Appendix for the variable definitions.

12 The daily data in this NYC test are “by date of diagnosis” which is presumably closer to the date the test is taken than the original (older) NYC data series (total confirmed by NYC DOH as of a certain date), or the NYS data series (total reported to the NYS system as of the date). New York State has total number of PCR tests by County which could be used to adjust the reported NYC data as if testing was done at a constant rate, but this adjustment is beyond the scope of this tutorial.

13 These figures in this paragraph are arbitrarily rounded to 1%. Also note that the precise calculation of a perfect seroprevalence test requires both the numerator and denominator be adjusted, i.e. (Σ*I* – *X*) / (*N – X*) where Σ*I* is the total cumulative infected. *S = S_pct_ N* and Σ*I = N* – *S*. For NYC *N ≅* 8,623,000. In numbers carrying extra precision to make the arithmetic clear (Σ*I* – *X*) / (*N* – *X*) ≅ 2599527/8468394 ≅ 28% for April 25th.

14 The statisticians and R programmers will recognize that the Model Run Output includes output of the summary () function applied on nlsLM() and lm() functions. The statistical estimates (i.e. standard errors, t-values and probabilities) have not been reviewed in this paper and their use may cause the user to draw non-factual conclusions about accuracy.

15 For purposes of this paper, which is an example demonstration of the effects of known sensitivity and specificity on the naive results, no calculation is made of the inaccuracy due to small sample size (i.e. confidence interval or distribution); and no adjustments are made for the known spread or distribution of specificity or sensitivity measurements. They exist and can be statistically computed if the information is available at the cost of added complexity.

